# Puumala orthohantavirus dysregulates hyaluronan metabolism in lung cells and correlates with disease severity and lung impairment

**DOI:** 10.64898/2026.03.10.26348053

**Authors:** Alfred Wennemo, Praveen Mathews Varghese, Urban Hellman, Nazar Beirag, Charlotta Andersson, Anders Blomberg, Johan Rasmuson, Clas Ahlm, Therese Thunberg, Annasara Lenman

## Abstract

Hantaviruses are globally distributed rodent-borne viruses that cause human diseases. In Europe, the Puumala orthohantavirus (PUUV) is the most prevalent hantavirus and causes a mild form of hemorrhagic fever with renal syndrome (HFRS), characterized by renal, hemorrhagic and pulmonary manifestations. To date, the mechanisms underlying pulmonary symptoms are still poorly understood, highlighting a significant gap in our knowledge of the disease. In this translational study, we investigated the role of hyaluronan (HA), an extracellular matrix glycosaminoglycan with a high water-retaining capacity, in pulmonary disease severity during PUUV infection and examined whether PUUV disrupts HA metabolism. We found that plasma HA levels increased during acute PUUV infection, normalized during convalescence, and correlated with disease severity. In lung tissue from fatal PUUV cases, HA accumulated extensively and was associated with disrupted alveolar architecture. Furthermore, in bronchoalveolar lavage fluid, high-molecular weight HA levels were elevated in patients with greater pulmonary involvement. In vitro, infecting a panel of human lung cell types, we found that PUUV infection altered HA homeostasis in a cell-type-specific manner. Infection induced an imbalance in HA metabolic pathways, with early upregulation of hyaluronan synthases followed by induction of HA degradation and receptor genes. Primary lung fibroblasts similarly showed increased HA production with pronounced donor variability in HA regulating gene expression not explained by infection levels. These findings identify dysregulated HA metabolism as a feature of PUUV infection and link pulmonary HA accumulation to disease severity, implicating HA as a potential biomarker and therapeutic target in hantavirus associated lung disease.

## INTRODUCTION

*Orthohantavirus* is a genus of highly pathogenic rodent-borne viruses within the Hantaviridae family. Infections caused by hantaviruses result in two distinct acute febrile diseases; hemorrhagic fever with renal syndrome (HFRS) in Europe and Asia and hantavirus cardiopulmonary syndrome (HPS) in the Americas [1]. Although recognized as distinct clinical entities, HFRS and HPS share several overlapping features. Both are characterized by a strong inflammatory response that affects the vascular endothelium, resulting in systemic disease [1]. While HFRS typically presents as a comparatively less severe illness, characterized by renal and cardiopulmonary manifestations [2–5]. HPS generally follows a more fulminant disease course, with rapid onset of respiratory failure, cardiogenic shock and severe hemorrhagic manifestations [6].

Hantaviruses usually circulate within one or a few closely related rodent species, geographically restricting their distribution [7]. Transmission to humans occurs via inhalation of aerosols contaminated with feces, saliva or urine from infected rodents. In Europe, the predominant hantavirus is *Puumala orthohantavirus* (PUUV) which causes a relatively mild version of HFRS (previously termed nephropathia epidemica) [8]. The clinical presentations of PUUV infections varies widely, ranging from subclinical or mild symptoms to multi-organ failure with fatal outcome [2–4]. Infections are characterized by high fever, headache, abdominal pain and nausea. These symptoms are non-specific to PUUV infections, making early diagnosis of the disease challenging. As the disease progresses, hallmark manifestations develop, including hemorrhagic, renal and pulmonary symptoms. While the hemorrhagic and renal complications have been extensively studied and well characterized, the mechanisms underlying the pulmonary pathology remain poorly understood [9]. These pulmonary symptoms are typically characterized by impaired gas diffusion, pulmonary oedema and thickening of connective tissue in the lungs, with reported symptoms varying from mild cough to acute respiratory distress syndrome (ARDS) [4, 10]. The lungs have been implicated as the primary site of PUUV infection, with infiltration of lymphocytes and presence of viral antigens in the lung vascular endothelium, epithelial and resident mononuclear cells [11–13]. Furthermore, *in vitro* studies indicate that bronchial, small-airway and alveolar epithelial cells are susceptible and permissive to PUUV [14]. However, key processes driving pulmonary pathology during PUUV infection have yet to be fully defined.

Hyaluronan (HA) is a non-sulphated glycosaminoglycan and an important structural component of the extracellular matrix, where it regulates cellular functions such as cell-matrix signaling, proliferation and migration [15]. In healthy tissue, HA is predominantly present as high-molecular weight HA (HMW-HA), where it exerts anti-inflammatory effects. In contrast, during inflammation, the turnover and degradation of HA increase, resulting in elevated concentrations of fragmented low-molecular weight HA (LMW-HA) [15]. These fragments have been shown to exert pro-inflammatory effects through activation of the NF-kB signaling pathway, further promoting the local inflammatory environment [15, 16]. In addition, HA has a remarkable capacity to retain water, enabling it to occupy large, hydrated volumes of up to 1000 times its molecular mass [15]. Consequently, HA plays an important role in tissue water homeostasis and contributes to oedema formation under pathological conditions. In the lungs, excessive HA accumulation is associated with pathogenesis of ARDS, where fluid builds up and impairs the gas exchange [16–18]. We previously showed that HA accumulates in the lungs of COVID-19 patients, filling the lungs with a clear liquid hydrogel that contribute to long-term lung impairment [17, 19]. Emerging evidence from other respiratory viral infections further implicates the role HA in respiratory disease [20, 21]. Together, these findings suggest a broader role for HA in respiratory viral disease and highlights its potential role of HA in the pulmonary pathogenesis of PUUV infections.

In this study, we investigated whether PUUV infection drives HA accumulation in the lungs of PUUV infected patients and contributes to the observed pulmonary pathogenesis. Using patient-derived lung tissue and bronchoalveolar lavage fluid (BALF), we demonstrate extensive HA accumulation in the lungs that correlates with increased pulmonary impairment. Furthermore, we show that PUUV infection dysregulates HA metabolism in alveolar epithelial cells and lung fibroblasts, leading to increased HA production and accumulation. Together, these findings identify HA dysregulation as a component of PUUV-associated lung pathology and highlight HA metabolism as a potential therapeutic target in PUUV infection.

## MATERIALS AND METHODS

### Description of the PUUV cohort

Fifty-four adult patients with serologically confirmed PUUV infection were prospectively enrolled in the study at the Department of Infectious Diseases, University Hospital of Umeå, Sweden. Clinical and laboratory parameters were recorded throughout the acute phase of the disease (0-2 weeks after disease onset), and at the convalescent phase visit (3-6 months after disease onset). Blood samples were collected at the corresponding time points and stored at −80 °C until analysis. Pulmonary involvement was studied in a subcohort of 15 participants, as previously described [10, 12]. These participants were investigated by a CT scan of the lungs and underwent bronchoscopy with collection of bronchoalveolar lavage fluid (BALF) during the acute phase of PUUV infection. The current study also includes three patients with fatal PUUV infection, whose clinical characteristics have been previously reported [2, 11].

### Definition of PUUV infection disease severity

Disease severity was categorized as either mild/moderate or severe based on to the clinical manifestations observed during the acute phase of PUUV infection according to a modified version of a previously described classification [22]. Patients were classified as having severe disease severity if they met two or more of the following clinical criteria: admission to the intensive care unit, moderate to severe hypotension (systolic blood pressure ≤ 90 mmHg requiring intravenous fluid therapy at any time during hospitalization), radiologically confirmed thrombosis, clinically significant bleeding, need of supplemental oxygen treatment, dialysis or platelet transfusion. All remaining patients were categorized as having mild/moderate disease.

### Measurement of hyaluronan concentration in patient plasma

Plasma from 54 patients during the acute and the convalescent phase of PUUV infection were analyzed for HA. Plasma samples from anonymous blood donors were included as healthy controls. HA concentrations were quantified using a competitive HA-binding protein-based ELISA-like assay (K-1200; Echelon Biosciences Inc., Salt Lake City, UT, USA), following the manufacturer’s instructions. Absorbance was measured on a Varioskan Flash Multimode Reader (Thermo Fisher Scientific, MA, USA) and HA concentrations were calculated by interpolating the absorbance values against a standard curve using a four-parameter logistic (4PL) regression model.

### Immunohistochemistry of lung tissue sections

Sections of distal lung tissue were obtained from autopsies of three deceased adult patients and from healthy regions of distal lung tissue from individuals undergoing lobectomy. Sections were deparaffinized in xylene and rehydrated gradually using decreasing ethanol solutions. Endogenous peroxidase activity was quenched with 3% H_2_O_2_ in methanol for 5 min, followed by PBS washes. Sections were blocked in 10 mg/ml bovine serum albumin (BSA) (Roche, Basel, Switzerland) in PBS for 30 min. To verify HA-specific staining, control sections were treated with 50 U/ml *Streptomyces* hyaluronidase (H3506; Sigma-Aldrich, St. Louis, MO, USA) for 4 h at 37 °C, then washed in PBS. All sections were subsequently incubated overnight at 4 °C with a biotin-conjugated hyaluronan-binding probe (1:40) (385911; Sigma-Aldrich, St. Louis, MO, USA). After PBS washes, VECTASTAIN® Elite ABC reagent (Vector Laboratories, Newark, CA, USA) was applied for 40 min, followed by PBS rinsing. HA-staining was visualized by incubating sections with 3,3′-diaminobenzidine (Vector Laboratories, Newark, CA, USA) for 5 min, followed by rinsing in tap water. Sections were counterstained with Mayer′s hematoxylin (Histolab Products AB, Askim, Sweden), rinsed with tap-water, dehydrated and mounted with DPX mounting media (Sigma-Aldrich, St. Louis, MO, USA). Lung sections of healthy and PUUV infected patients were similarly stained for CD45 to visualize lymphocytes in the tissue. For this, the primary CD45-binding antibody (760-2505; Ventana Medical Systems, Tucson, AZ, USA) and the DAB-based detection kit (760-500; Ventana Medical Systems, Tucson, AZ, USA) were used according to manufactures instructions. Sections were counterstained with Hematoxylin II (Ventana Medical Systems, Tucson, AZ, USA) followed by Bluing Reagent (Roche, Basel, Switzerland) and mounted. Slides were scanned using a PANNORAMIC 250 Flash III scanner (3DHISTECH Ltd., Budapest, Hungary) with PANNORAMIC Scanner Software v3.03, and images were analyzed in QuPath software version 0.5.1 [23].

### Hyaluronan size fractionation and measurement of hyaluronan concentration in bronchoalveolar lavage fluid

Bronchoalveolar lavage fluid (BALF) was collected from 15 patients with acute-phase PUUV infection and 13 healthy controls using flexible video bronchoscopy. Three 60 mL aliquots of PBS were infused and retrieved from the right middle lobe, pooled, and stored at −80 °C after the procedure [12]. Total HA from BALF was extracted and size-fractioned from BALF by ion exclusion chromatography using a modified version of a previously described method [24]. In short, proteins crosslinked to HA were digested by treating samples with 1.8 U/ml Proteinase K (Thermo Fisher Scientific, Waltham, MA, USA) in the presence of 5 mM deferoxamine mesylate (Sigma-Aldrich, St. Louis, MO, USA) to prevent iron-associated HA depolymerization. Samples were incubated overnight at 55 °C, after which Proteinase K was inactivated by heating at 90 °C for 5 min. Any aggregated HA fragments were dissolved by shaking at 800 rpm for 30 min. Samples were then loaded on anion-exchange columns (Thermo Fisher Scientific, Waltham, MA, USA) pre-equilibrated with 0.1 M NaCl and centrifuged at 2000 x g for 5 min. Columns were washed three times with PBS under the same centrifugation conditions. Bound HA was stepwise eluted with 0.46 M and 0.7 M NaCl by centrifugation at 400 x g for 15 min. Eluted fractions were transferred to 3-kDa molecular-weight cut-off filters (Millipore, Burlington, MA, USA) prewashed with PBS. Each fraction was washed three times with PBS at 14 000 x g for 5 min to remove excess NaCl, the retained HA was eluted by inverting the filters and centrifuging at 1 000 x g for 2 min. Final volumes were adjusted to 250 μl and HA concentrations were quantified using an ELISA-like HA-concentration measurement kit (K-1200; Echelon Biosciences Inc., Salt Lake City, UT, USA) as described under *“Measurement of hyaluronan concentration in patient plasma”*.

### Cells lines and reagents

Human lung fibroblasts MRC-5 (CCL-171™, ATCC, Manassas, VA, USA), lung carcinoma alveolar epithelial cells A549 (CCL-185™, ATCC, Manassas, VA, USA), lung bronchial epithelial cells BEAS-2B (CRL-3588™, ATCC, Manassas, VA, USA) and African green monkey kidney epithelial cells Vero E6 (CRL-1586™, ATCC, Manassas, VA, USA) were cultured in DMEM supplemented with 20 mM HEPES, 100 U/mL Penicillin-Streptomycin and 10 % FBS. Modified A549 cells expressing the IFN antagonist PIV5 V protein (A549/PIV5-V, a kind gift from Dr Catherine Adamson, University of St Andrews, UK), were maintained in the same growth media supplemented with 2 μg/mL puromycin (Gibco, Thermo Fisher Scientific, Waltham, MA, USA). Human lung microvascular endothelial cells HULEC-5a (CRL-3244™, American Type Culture Collection, Manassas, VA, USA) were cultured in MCDB 131 medium (Sigma-Aldrich, St. Louis, MO, USA) supplemented with 10 ng/mL human epidermal growth factor (Sigma-Aldrich, St. Louis, MO, USA), 1 μg/mL hydrocortisone (Sigma-Aldrich, St. Louis, MO, USA), 100 U/mL Penicillin-Streptomycin and 10% FBS. All cell lines were maintained in a humidified incubator at 37 °C with 5 % CO_2_ and routinely tested negative for mycoplasma.

### Verification of A549/PIV5-V STAT1-knockdown

Human lung carcinoma alveolar epithelial A549 cells and the modified A549/PIV5-V cell line were seeded at a density of 1,000,000 cells per well in 6-well plates (VWR, Radnor, PA, USA). Cells were incubated for 24h at 37 °C with 5% CO_2_ to allow monolayer formation. The growth media was removed, and cells were lysed in RIPA buffer (50 mM TRIS, 150 mM NaCl, 1,5% (v/v) Triton X-100, 0.5% (w/v) Sodium deoxycholate and 0.1% (w/v) SDS). Cell lysates were stored at −20 °C until analysis.

Proteins in cell lysate were separated by sodium dodecyl sulfate–polyacrylamide gel electrophoresis (SDS–PAGE) using precast 4–12% Bis-Tris Mini Protein Gels (Invitrogen, Carlsbad, CA, USA) and transferred onto a PVDF membrane (Millipore, Burlington, MA, USA). The membrane was blocked with 5% (w/v) skimmed milk in PBS, followed by overnight incubation with monoclonal anti-STAT1 antibody (1:500) (AHO0832, Invitrogen, Carlsbad, CA, USA) and β-actin antibody (1:1000) (MA5-15739, Invitrogen, Carlsbad, CA, USA). After washing with PBS-Tween (Medicago, Uppsala, Sweden), the membrane was incubated with HRP-conjugated anti-mouse secondary antibody (1:5000) (Invitrogen, Carlsbad, CA, USA). Chemiluminescent substrate (Thermo Fisher Scientific, Waltham, MA, USA) was used for detection, and bands were visualized using the Amersham Imager 580 (GE Healthcare Life Sciences, Uppsala, Sweden).

### Puumala orthohantavirus propagation and titration

Confluent Vero E6 cells were inoculated with PUUV (PUUV strain Kazan, adapted to cell culture as previous described [25]) at a multiplicity of infection (MOI) of 0.002 for 2h at 37 °C with 5% CO_2_. Following adsorption, fresh DMEM supplemented with 20 mM HEPES, 100 U/ml Penicillin-Streptomycin, and 2% FBS was added. Cell culture supernatants were harvested at 7 days post-infection (dpi), clarified by centrifugation at 2000 x g for 5 min, and stored at 4 °C. A second harvest was performed at 14 dpi and clarified as described above. Virus-containing supernatants were purified by ultracentrifugation through a 30% (w/v) sucrose cushion (30% sucrose, 20 mM HEPES, 150 mM NaCl) using an Optima L-80 XP ultracentrifuge (Beckman Coulter) with a SW 32 rotor at 4 °C for 2 h. Viral pellets were resuspended in DMEM and stored at −80 °C until use.

Viral titers were determined by focus-forming assay. Vero E6 cells were seeded at a density of 25,000 cells per well in black 96-well optical-bottom plates (Greiner Bio-One International GmbH, Kremsmünster, Austria) and incubated overnight at 37 °C with 5% CO_2_. Cells were washed once with DMEM supplemented with 20 mM HEPES and 100 U/ml Penicillin-Streptomycin, then infected with a 10-fold serial dilution of PUUV. After 2 h adsorption at 37°C, the inoculum was removed and growth media supplemented with 2% FBS was added to the cells. Infection was allowed to progress for 22-24 h, after which cells were fixated with 4% paraformaldehyde (Thermo Fisher Scientific, Waltham, MA, USA) for 10 min. Cells were subsequently stained as described under “*Immunofluorescent staining*“, and PUUV antigen-positive cell foci were quantified.

### Puumala orthohantavirus infection of human lung cells

Human cell lines representing the different cell types of the lungs; MRC-5, A549, A549/PIV5-V, BEAS-2B, and HULEC-5a cells were seeded at a density of 25,000 cells per well in black 96-well optical-bottom plates for infection visualization and at 150,000 cells per well in 24-well plates (VWR, Radnor, PA, USA) for gene-expression analysis. Cells were incubated for 16-20 h at 37 °C with 5% CO_2_ to allow monolayer formation. Prior to infection, cells were washed with DMEM supplemented with 20 mM HEPES and 100 U/mL Penicillin-Streptomycin. PUUV was added at a (MOI) of 2.5 to all cell lines except HULEC-5a cells, which were infected at a MOI of 0.25. Virus adsorption was allowed for 2 h at 37 °C with 5% CO_2_. Unbound viruses were removed by washing and cells were returned to their respective growth media supplemented with 2% FBS and incubated at 37 °C in 5% CO_2_. Progression of infection was monitored for up to 5 days (120 hours post-infection (hpi)). For assessment of viral propagation, cells were fixed at 48, 72, 96 and 120 hpi with 4% paraformaldehyde (Thermo Fisher Scientific, Waltham, MA, USA) for 10 min. For quantification of released HA, cell culture media from the same plates were collected at the specified time points and stored at −20 °C until analysis. To analyze infection-induced changes in gene expression upon infection, cells were lysed at 24, 48, 72 and 96 hpi using lysis buffer (740984; Macherey-Nagel GmbH & Co. KG, Düren, Germany), and lysates were stored at −20 °C until further processing.

### Immunofluorescent staining

Infected cells were quantified by immunofluorescent detection of viral antigens in the cytosol. Fixed cells were permeabilized with ice-cold methanol for 10 min at −20 °C and subsequently blocked with 2% BSA in PBS for 1 h. PUUV nucleocapsid protein was detected using a polyclonal primary antibody (1:500) (NR-9675; BEI Resources, Manassas, VA, USA) followed by an Alexa Fluor 488-conjugated secondary antibody (1:1000) (A-11008; Invitrogen, Carlsbad, CA, USA) diluted in 2% BSA in PBS. Cell nuclei were counterstained with Hoechst 13342 (1:10000) (Thermo Fisher Scientific, Waltham, MA, USA). Plates were imaged using a Cytation 5 Cell Imaging Multimode Reader (Agilent BioTek, Winooski, VT, USA), and images were analyzed in Gen5 software v3.2 to quantify total and infected cell counts.

### Measurement of hyaluronan concentrations in cell growth media

Concentration of HA in the cell culture media were quantified using a sandwich HA-binding protein-based ELISA-like assay (DY3614; R&D Systems, Minneapolis, MN, USA) according to the manufacturer’s instructions. Absorbance was measured using a Varioskan Flash reader (Thermo Fisher Scientific, MA, USA), and concentrations were calculated by 4-parameter logistic regression against a standard. HA levels in infected samples were normalized to those of time-matched uninfected controls.

### Measurement of gene expression with quantitative PCR

Total RNA was extracted from cell lysates using the NucleoSpin RNA Plus kit (Macherey-Nagel GmbH & Co. KG, Düren, Germany) and 100 ng of RNA was reverse-transcribed to cDNA using the High-capacity cDNA Reverse transcription Kit (Applied Biosystems, Thermo Fisher Scientific, Waltham, MA, USA). Gene expression of *HAS1*, *HAS2*, *HAS3*, *HYAL1*, *HYAL2* and *CD44* was measured by quantitative PCR using qPCRBIO SyGreen® Mix Lo-ROX (PCR Biosystems) and the QuantiTect primer assays (QIAGEN, Hilden, Germany): *HAS1* (QT02588509), *HAS2* (QT00027510), *HAS3* (QT00014903), *HYAL1* (QT01673413), *HYAL2* (QT00013363), and *CD44* (QT00998333). Reactions were run on the QuantStudio 5 system (Applied Biosystems, Thermo Fisher Scientific, Waltham, MA, USA). Gene expression levels were normalized to housekeeping B-actin (QT01680476; QIAGEN, Hilden, Germany), and expression in infected samples was normalized to time-matched uninfected controls and reported as relative expression.

### Isolation of primary lung fibroblasts from distal lung tissue

Primary human fibroblasts were isolated from distal lung tissue obtained from individuals undergoing thoracic surgery at the University Hospital, Umeå, Sweden. Tissue pieces (≤5 mm) were placed onto tissue culture-treated dishes (Corning Incorporated, Corning, NY, USA) precoated with Earle’s Balanced Salt Solution (Sigma-Aldrich, St. Louis, MO, USA) supplemented with 3 µg/ml collagen (Sigma-Aldrich, St. Louis, MO, USA), 1µg/ml BSA (Sigma-Aldrich, St. Louis, MO, USA) and 1µg/ml fibronectin (Sigma-Aldrich, St. Louis, MO, USA). Fibroblasts were maintained in Dulbecco’s Modified Eagle Medium (DMEM) (Sigma-Aldrich, St. Louis, MO, USA) supplemented with 20 mM HEPES, 100 U/ml Penicillin-Streptomycin (Gibco, Thermo Fisher Scientific, Waltham, MA, USA) and 10% fetal bovine serum (FBS) (Cytiva, Marlborough, MA, USA). Cells were kept in a humidified incubator at 37 °C with 5% CO_2_. Primary cells from five different donors were used at a passage number between 3-5. The fibroblasts were seeded and infected as described under “*Puumala orthohantavirus infection of lung cells*” at a MOI of 2.5 and infection was followed for four days.

### Statistical analysis

Statistical analysis was performed using GraphPad prism 10 (version 10.1.1). Patient groups were compared using Mann-Whitney U test. Correlation between HA concentration and clinical parameters were assessed with simple linear regression analysis. Infection levels were compared using unpaired t-tests. HA levels in culture media were analyzed using multiple unpaired t-tests with Holm–Šídák correction for multiple comparisons. Differences in gene expression were analyzed using paired t-tests, comparing the housekeeping-normalized gene expression in infected samples to time-matched uninfected controls. Statistical significance is denoted as follows: \**P* < 0.05, \*\**P* < 0.01, \*\*\**P* < 0.001; ns, not significant.

### Ethical approval

The study was performed according to the Declaration of Helsinki and approved by the Swedish Ethical Review Authority (Dnr. 04-113M & Dnr. 07-162M for the patient cohort and Dnr. 2021-00317 for collecting primary lung cells). Patients were included after informed consent.

### Data availability

All data produced in the present study are available upon reasonable request to the authors, except for patient data which cannot be made publicly available according to Swedish data protection laws.

## RESULTS

### Systemic hyaluronan levels increase during the acute phase of PUUV infection

Given the previously reported relationship between HA and COVID-19 [17, 19], we here set out to investigate whether systemic HA levels are similarly altered in patients during PUUV infection. To explore this, we quantified plasma HA concentrations in a well-characterized patient cohort and assessed their association with disease severity. Plasma samples were obtained from 54 patients during the acute phase (0-2 weeks after disease onset) and the convalescent phase (3-6 months after disease onset) of infection. The demography and clinical characteristics of the study participants are presented in Table 1. Participants ranged from 23 to 82 years of age, and the cohort consisted of more females (n=30) than males (n=24). Most patients were previously healthy. Among those with comorbidities, cardiovascular disease, including hypertension, heart failure, arrhythmias, or prior stroke, was the most common. A total of 7% had a documented chronic lung disease (asthma or chronic obstructive pulmonary disease), and 9% had type 2 diabetes. Smoking was reported in 20% of patients; however, smoking status was missing for a large proportion of the cohort (35%). The study was conducted at Umeå University Hospital, where most patients (76%) required inpatient care, including three who required treatment at the intensive care unit.

**Table 1.**
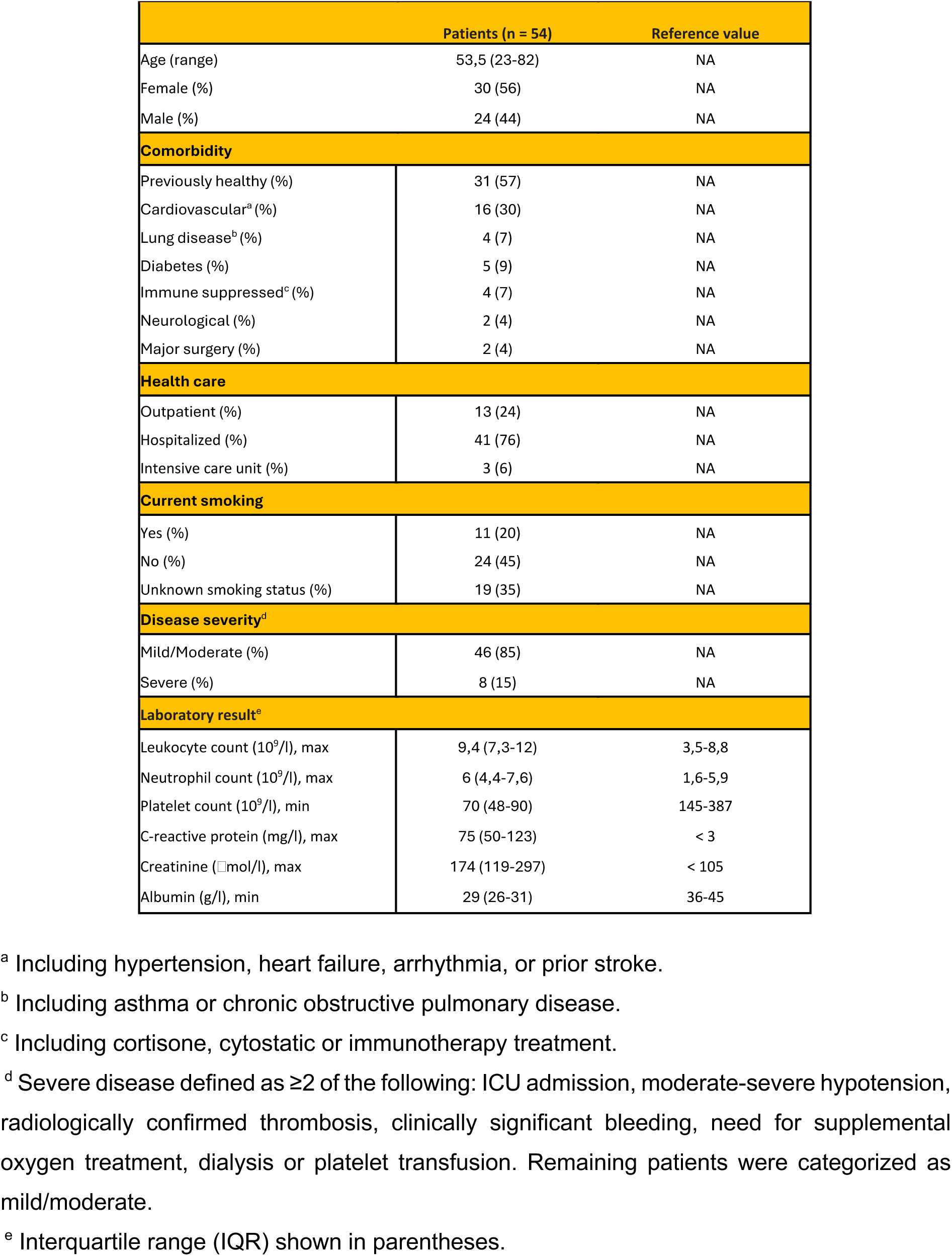
Demography and characteristics of PUUV patients.

Plasma HA concentrations were quantified using ELISA and compared with healthy controls (Fig. 1A). During the acute phase of PUUV infection, systemic HA levels were elevated in patients with both mild/moderate (P < 0.001) and severe (P < 0.001) disease compared with healthy controls (Fig. 1B). Notably, patients with severe disease exhibited markedly higher HA concentrations than those with mild/moderate disease (P < 0.05). By the time patients reached convalescence, HA levels in both groups had returned to levels comparable to those of healthy controls (Fig. 1C). Together, these results demonstrate that systemic HA is markedly elevated during acute PUUV infection, increases in proportion to disease severity, and returns to baseline during disease recovery.

**Figure 1.**
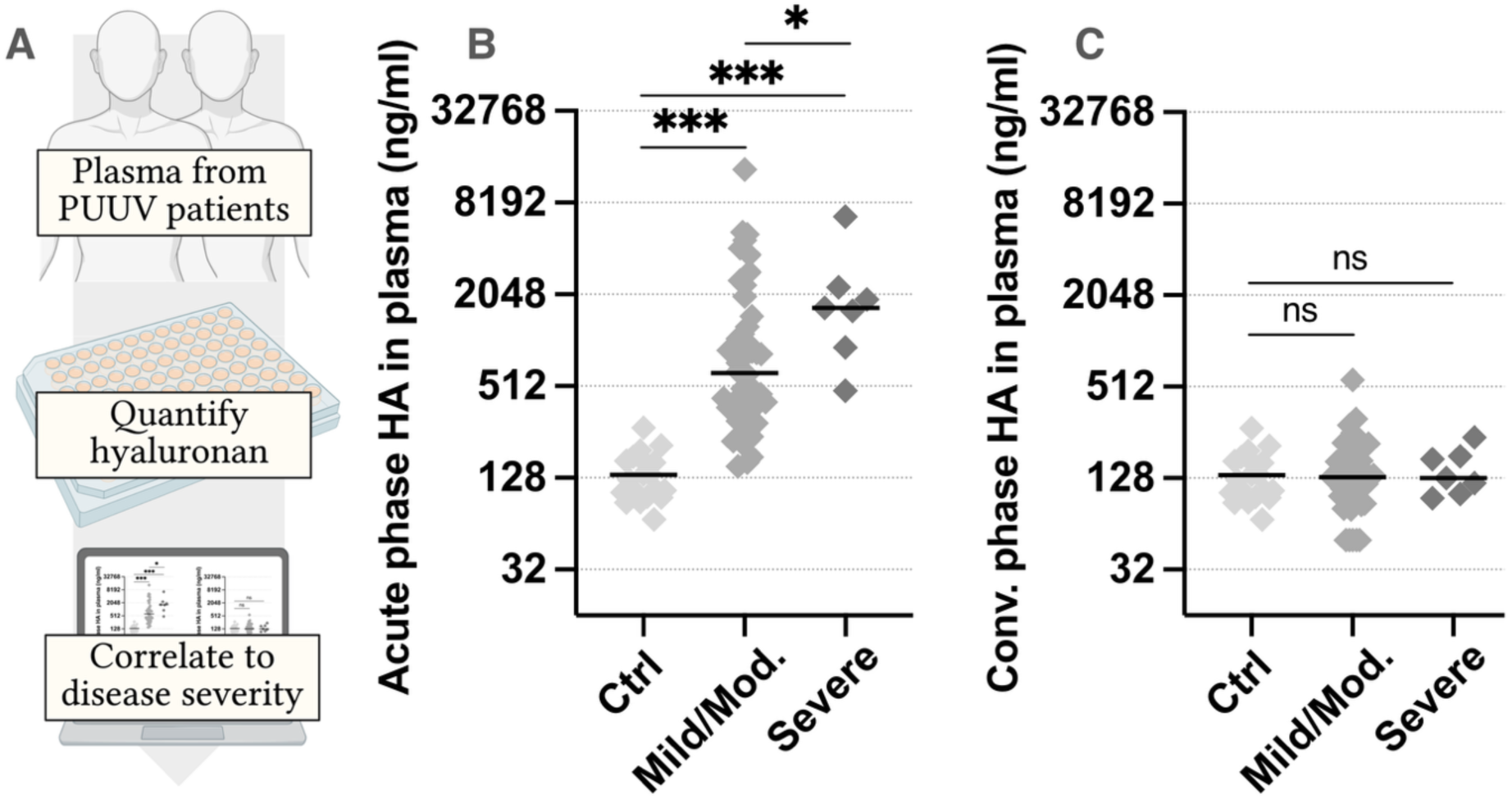
Elevated hyaluronan (HA) levels in plasma from Puumala orthohantavirus (PUUV) infected patients correlate with disease severity. (A) Schematic overview of plasma collection and HA analysis. (B–C) Plasma HA concentrations during the (B) acute phase (0–2 weeks after disease onset) and (C) convalescent phase (3-6 months after disease onset) of the disease. HA concentrations were quantified by ELISA and compared to healthy controls. Each square represents one individual, and horizontal lines indicate the median. Patients were classified with severe disease if they met two or more of the following clinical criteria: admission to the intensive care unit, moderate to severe hypotension (systolic blood pressure ≤ 90 mmHg requiring intravenous fluid therapy at any time during hospitalization), radiologically confirmed thrombosis, clinically significant bleeding, need for supplemental oxygen treatment, dialysis or platelet transfusion. All other patients were classified as mild/moderate disease. Statistical significance was assessed using the Mann-Whitney U test (\**P* < 0.05, \*\**P* < 0.01, and \*\*\**P* < 0.001; ns, not significant).

### Hyaluronan accumulates in the lungs in PUUV infection

Given the systemic HA increase observed in patients with PUUV infection, we next examined whether HA also accumulates locally in the lungs during acute infection. We performed histochemical analysis of lung tissue from deceased patients and compared HA distribution and tissue morphology with healthy lung sections. There were striking morphological differences between patients and healthy lung tissue (Fig. 2A). Healthy lungs displayed well-defined, air-filled alveoli with HA confined to the interalveolar walls and perivascular regions. In contrast, lungs from deceased patients exhibited denser morphology with excessive accumulation of HA (Fig. 2A). Detailed examinations further revealed distinct regions representing progressive stages of alveolar disruption, displaying alveoli partly filled with HA- rich exudate and alveoli fully packed with HA, showing complete loss of alveolar architecture. The alveoli disruption was accompanied by thickened alveolar walls and progressive depletion of normal wall-associated HA matrix. Enzymatic pretreatment of lung sections with hyaluronidase abolished all HA staining, confirming the specificity of the histochemical staining (Fig. 2B). Moreover, increased immune cell infiltration was observed in the infected tissue (Fig. S1A), with up to a 3.4-fold increase in leukocyte infiltration compared to healthy controls (Fig S1B-C).

**Figure 2.**
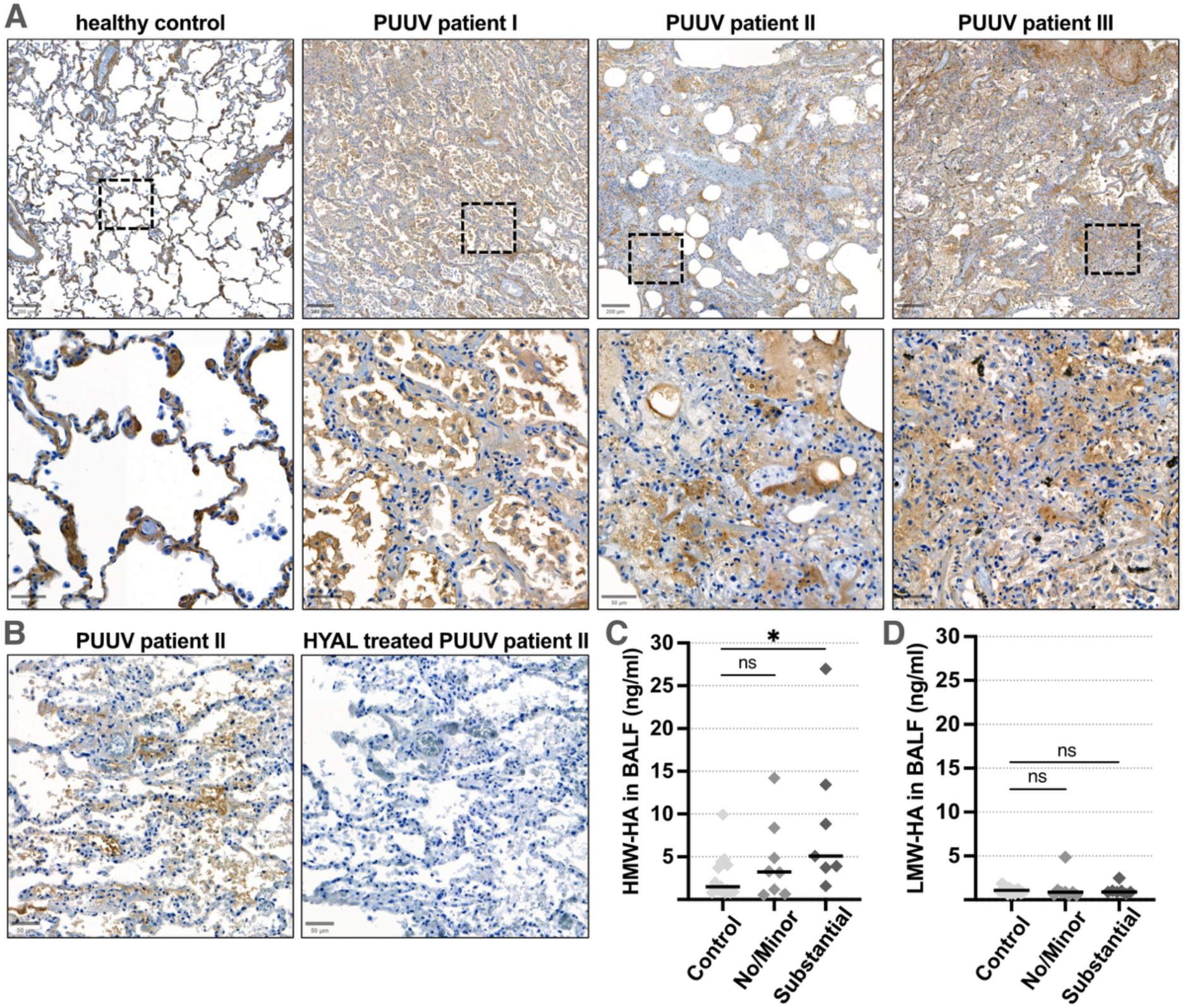
Hyaluronan (HA) accumulates in lung tissue and bronchoalveolar lavage fluid from Puumala orthohantavirus (PUUV) infected patients. (A) Lung tissue sections from healthy controls and deceased PUUV infected patients (n = 3) were stained for HA using a hyaluronan-binding protein (brown) and counterstained with hematoxylin to visualize nuclei (blue). The upper panel shows a representative overview of the healthy tissue and tissue from the three deceased PUUV-infected patients. Scale bar, 200 µm. The lower panel displays higher-magnification images of indicated regions from the same tissue sections, highlighting the altered morphology in the PUUV-infected lungs. Scale bar, 50µm (B) Representative PUUV-infected lung section before and after hyaluronidase (HYAL) treatment, showing the complete loss of HA signal and confirming staining specificity. Scale bar, 50µm. (C–D) Size-fractionated HA isolated from bronchoalveolar lavage fluid (BALF) of PUUV infected patients and healthy controls, separated into (C) high–molecular weight (HMW) and (D) low–molecular weight (LMW) HA fractions by anion-exchange chromatography. Each square represents one individual, and horizontal lines indicate the median. Patients were classified as having substantial pulmonary involvement if they met two or more of the following pulmonary manifestations: abnormal chest CT findings, respiratory symptoms or need for supplementary oxygen. All other patients were classified as having no/minor pulmonary involvement. Statistical significance was assessed using the Mann-Whitney U test (*P < 0.05; ns, not significant).

To determine whether pulmonary HA accumulation correlated with pulmonary involvement, and to characterize the molecular size distribution of HA, we quantified HMW- and LMW-HA in bronchoalveolar lavage fluid (BALF) from patients and healthy controls. Patients with PUUV infection were categorized as having substantial pulmonary involvement if they met at least two of the following criteria: abnormal chest CT findings, respiratory symptoms or requirement for supplementary oxygen. All remaining patients were categorized as having no or minor pulmonary involvement. HMW-HA concentrations were elevated in patients with substantial pulmonary involvement (P < 0.05) and displayed a similar, though non-significant, increase in patients with no/minor involvement (Fig. 2C). In contrast, LMW-HA levels remained unchanged across both groups compared to healthy controls (Fig. 2D). Although HMW-HA levels were elevated in patients with pulmonary involvement, no significant correlation was observed between HMW-HA concentrations and pulmonary diffusion capacity in infected patients (Fig. S2). This analysis was limited by the relatively small sample size.

Together, these findings show that HA accumulates extensively in the lungs of PUUV infected patients and that elevated HMW-HA in BALF is associated with greater pulmonary involvement, indicating a role for HA in the pulmonary manifestations of acute PUUV infection.

### Alveolar epithelial and fibroblast cells accumulate hyaluronan upon PUUV infection

Having observed extensive HA deposits in the lungs of patients, we next investigated whether PUUV infection directly alters HA metabolism in human lung cells. We infected a panel of lung-derived cell lines representing the major pulmonary cell types, including alveolar epithelial (A549), bronchial epithelial (BEAS-2B), lung fibroblasts (MRC-5) and lung endothelial (HULEC-5a) cells (Fig. 3A). To examine the contribution of type I and type II interferon responses, we also included an A549-derived cell line expressing the parainfluenza virus 5 (PIV5) IFN antagonist V protein (A549/PIV5-V), which suppresses STAT1-dependent antiviral responses through proteasomal degradation of STAT1. The absence of STAT1 in A549/PIV5-V cells was confirmed by western blot, showing markedly reduced STAT1 levels compared to A549 cells, confirming the STAT1 knockdown (Fig. S3). To characterize the permissiveness of each cell type, we quantified infection over a five-day time course. The cell lines displayed distinct infection profiles, with A549, BEAS-2B and HULEC-5a cells showing a gradual decline in infection levels over time (Fig. 3B-D). MRC-5 cells maintained stable infection levels, indicating a limited spread (Fig. 3E). In contrast, A549/PIV5-V exhibited a progressive increase in infection over time, demonstrating efficient viral propagation (Fig. 3F). Of note, HULEC-5a cells required a tenfold lower MOI to achieve infection levels comparable to those observed in the other cell lines, consistent with endothelial cells being the main target of PUUV *in vivo*. Despite variability in replication dynamics, all cell types produced infectious progeny viruses (data not shown).

**Figure 3.**
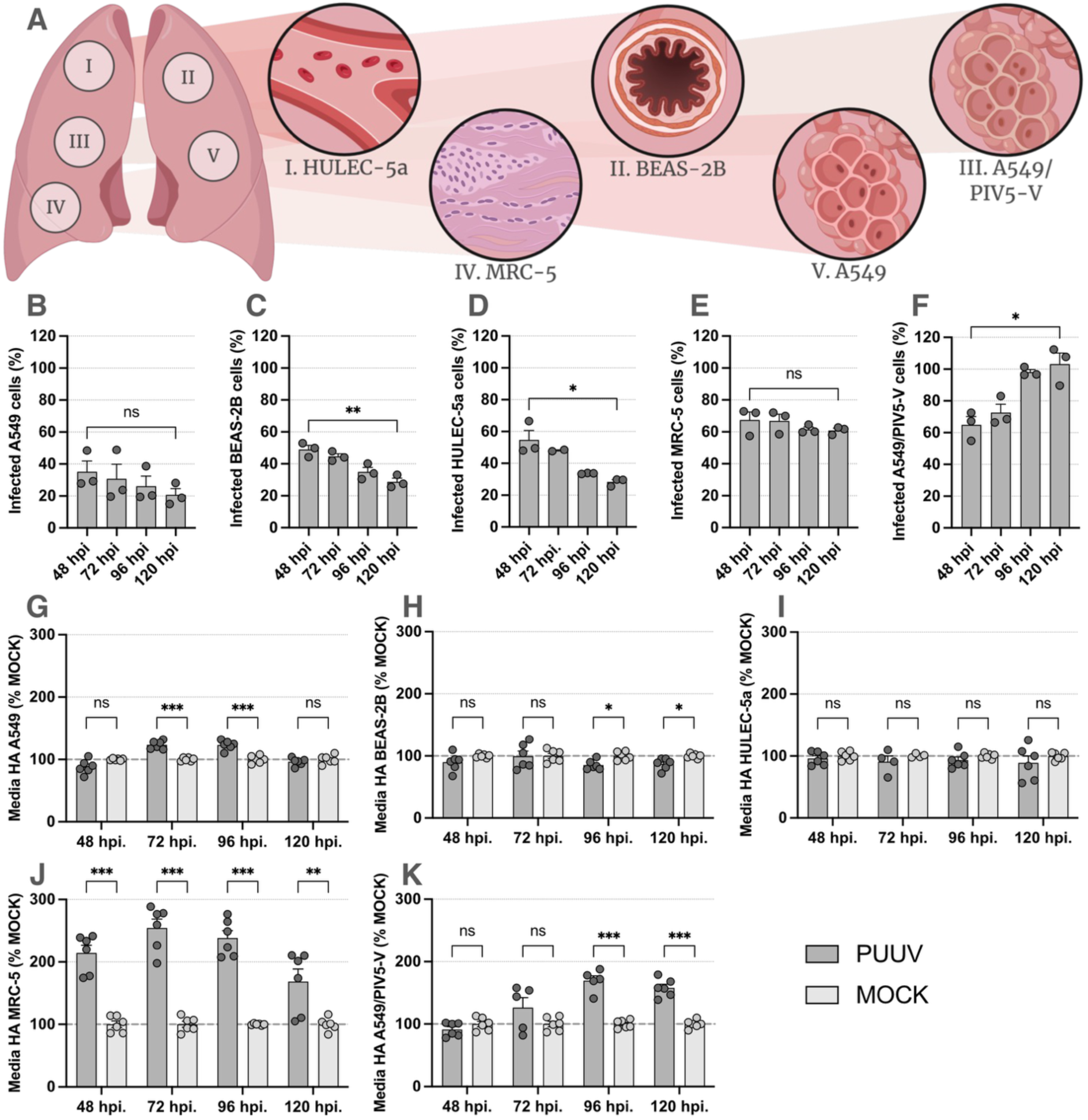
Puumala orthohantavirus (PUUV) infection induces cell type-dependent hyaluronan (HA) accumulation in human lung cells. (A) Schematic overview of the human lung cell lines used in the study. (B–F) PUUV infection kinetics over five days, quantified as the percentage of infected cells by immunofluorescence staining for PUUV nucleocapsid protein with nuclear counterstaining in (B) A549 alveolar epithelial cells, (C) BEAS-2B bronchial epithelial cells, (D) HULEC-5a microvascular lung endothelial cells, (E) MRC-5 lung fibroblasts, and (F) A549/PIV5-V cells with impaired STAT1 signaling. Statistical significance was calculated by unpaired t-test (*P < 0.05, **P < 0.01; ns, not significant). (G–K) HA levels in culture media supernatants collected at the indicated time points from PUUV-infected (G) A549, (H) BEAS-2B, (I) HULEC-5a, (J) MRC-5, and (K) A549/PIV5-V cells. HA concentrations were measured by ELISA and are shown as percentage of mock-infected controls. Data represents three independent experiments with duplicate technical replicates per experiment. Bars show mean ± SEM. Statistical significance was assessed using multiple unpaired t-tests with Holm–Šídák correction for multiple comparisons (*P < 0.05, **P < 0.01, ***P < 0.001; ns, not significant).

HA concentrations in culture media were quantified by ELISA throughout the infection and compared to time-matched uninfected controls. Interestingly, HA accumulation occurred only in a subset of cell lines, revealing a cell-type specific effect of PUUV infection on HA metabolism. Elevated HA levels were detected in A549, MRC-5 and A549/PIV5-V cells, with peak accumulation between 72 and 96 hpi (Fig. 3G, J, K). MRC-5 cells showed the greatest response, reaching 254% of control levels, while A549 and A549/PIV5-V reached 123% and 170%, respectively. In contrast, BEAS-2B and HULEC-5a cells did not accumulate HA, BEAS-2B even exhibited reduced HA levels late in infection (Fig. 3H-I). Taken together, these data suggest a cell-type specific response to PUUV infection, resulting in either increased or depleted HA levels in the culture media.

Collectively, these findings show that PUUV infection alters HA metabolism in a cell-type-specific manner independently of STAT1-mediated interferon signaling, with alveolar epithelial cells and fibroblasts serving as the primary sources of HA accumulation, thereby further implicating HA in the pulmonary pathogenesis of PUUV infection.

### PUUV infection dysregulates HA synthesis and degradation pathways

Given the PUUV-induced alterations in HA production observed across lung cell types, we next examined how infection alters the expression of key genes involved in HA metabolism. Transcript levels of HA-synthesizing enzymes (hyaluronan synthase; *HAS1-3)*, HA-degrading enzymes (hyaluronidase; *HYAL1-2*) and the primary HA receptor (*CD44*) were quantified by qPCR over a four-day infection time course and compared with time-matched uninfected controls (Fig. 4A). Quantified expression of the hyaluronan synthases revealed upregulation of synthases in all cell lines, even those without detectable HA accumulation, however *HAS1* expression was undetectable in all cell lines (Fig. 4B-F). In A549 cells, *HAS2* was upregulated already at 48 hpi and remained elevated throughout infection, whereas *HAS3* expression gradually declined (Fig. 4B). MRC-5 and A549/PIV5-V showed upregulation of both *HAS2* and *HAS3*, though with different temporal patterns. In MRC-5 cells, both *HAS2* and *HAS3* were initially upregulated, but while *HAS2* expression later returned to baseline, *HAS3* expression remained elevated (Fig. 4E). In A549/PIV5-V cells, *HAS2* expression gradually increased across the infection whilst *HAS3* displayed early, sustained upregulation (Fig. 4F). *HAS2* expression was not detected in BEAS-2B or HULEC-5a, however both cell types displayed induction of *HAS3*, with BEAS-2B showing a sustained response and HULEC-5a exhibiting a weaker, transient increase (Fig. 4C-D). Notably, the largest relative increase in hyaluronan synthase expression was observed in MRC-5 cells, which also exhibited the most pronounced HA accumulation (Fig. 3J, 4E).

**Figure 4.**
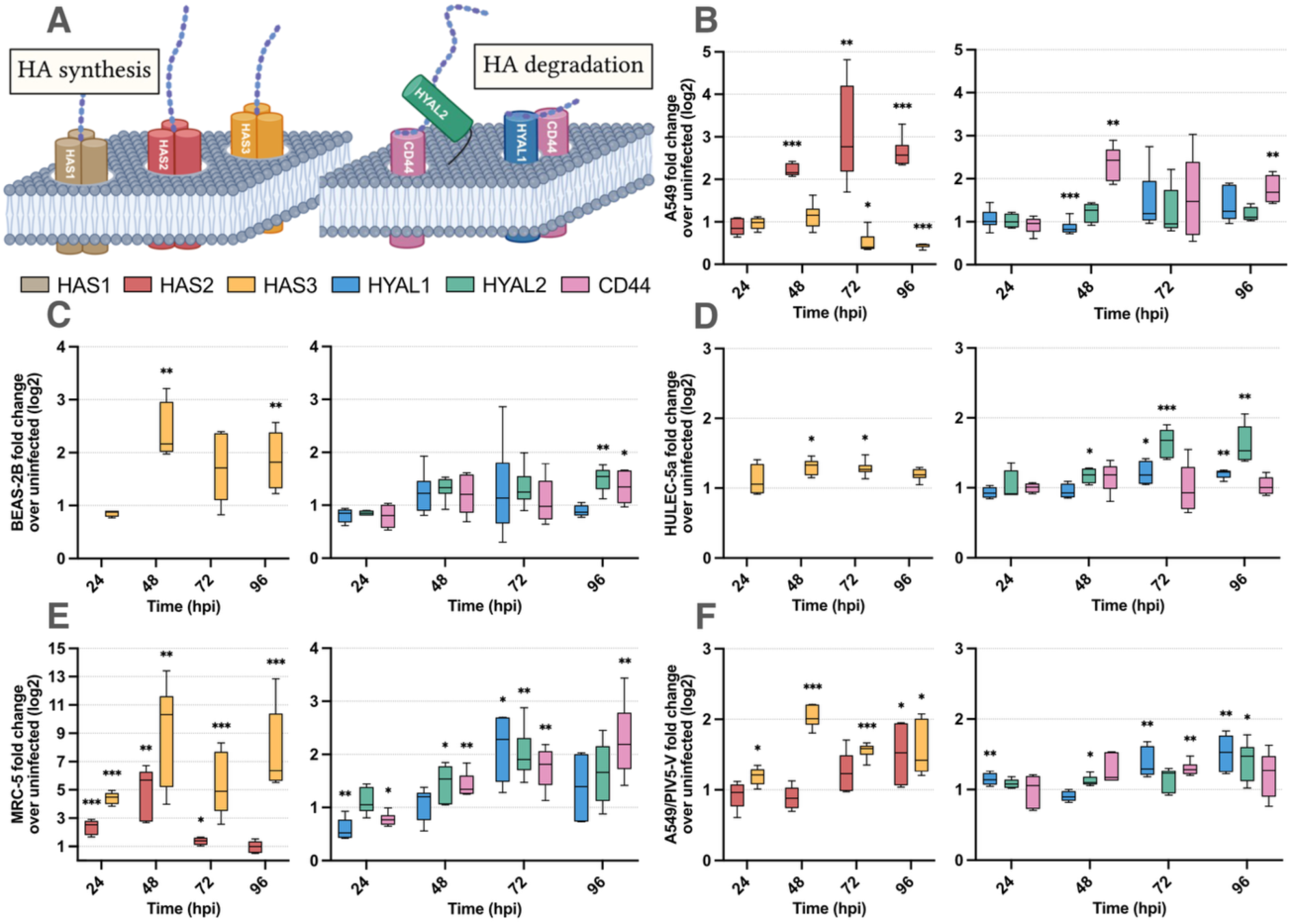
Puumala orthohantavirus (PUUV) infection of human lung cells causes dysregulation of genes involved in hyaluronan (HA) metabolism and signaling. (A) Schematic overview of genes involved in HA synthesis (*HAS1–3*), HA degradation (*HYAL1–2*), and HA signaling (*CD44*). (B–F) Expression of HA-regulating genes in (B) A549, (C) BEAS-2B, (D) HULEC-5a, (E) MRC-5, and (F) A549/PIV5-V cells, determined by quantitative PCR at the indicated time points. Expression levels were normalized to β-actin and are shown as fold change relative to uninfected controls. Data represents three independent experiments with duplicate technical replicates per experiment. Results are displayed as box-and-whisker plots, where the box indicates the 25th to 75th percentiles (interquartile range), the line within the box represents the median, and the whiskers denote the minimum and maximum values. Statistical significance was assessed using paired t-tests, comparing the expression values to the respective time-matched uninfected controls (*P < 0.05, **P < 0.01, ***P < 0.001).

Expression of HA-degrading enzymes showed an overall delayed and less pronounced response compared to the HA synthases. In A549 cells, *HYAL1* was downregulated during early infection and then gradually increased, whereas *CD44* was upregulated early and remained elevated (Fig. 4B). MRC-5 cells exhibited a similar late-stage upregulation of *HYAL1*, *HYAL2* and *CD44* (Fig. 4E), while A549/PIV5-V cells showed induction of *HYAL1* and *HYAL2* only (Fig. 4F). A comparable trend was observed in BEAS-2B and HULEC-5a cells, where BEAS-2B exhibited increased *HYAL2* and *CD44* expression (Fig. 4C), and HULEC-5a displayed upregulation of both *HYAL1* and *HYAL2* (Fig. 4D).

Together, these findings show that PUUV infection reprograms HA metabolic pathways in a cell-type-specific manner, creating an imbalance between HA synthesis and degradation.

### Infection of primary fibroblasts reveal donor-specific PUUV susceptibility and hyaluronan responses

The cell-line experiments revealed pronounced cell-type-specific differences in HA dysregulation during PUUV infection. Given the substantial inter-individual variability in pulmonary involvement observed among PUUV infected patients, we next examined whether primary human lung cells exhibit similar heterogeneity. Given that MRC-5 cells showed the most pronounced HA response, we isolated primary human lung fibroblasts (HLFs) from five individual donors and infected them with PUUV to assess donor-dependent infection kinetics and HA production (Fig. 5A).

**Figure 5.**
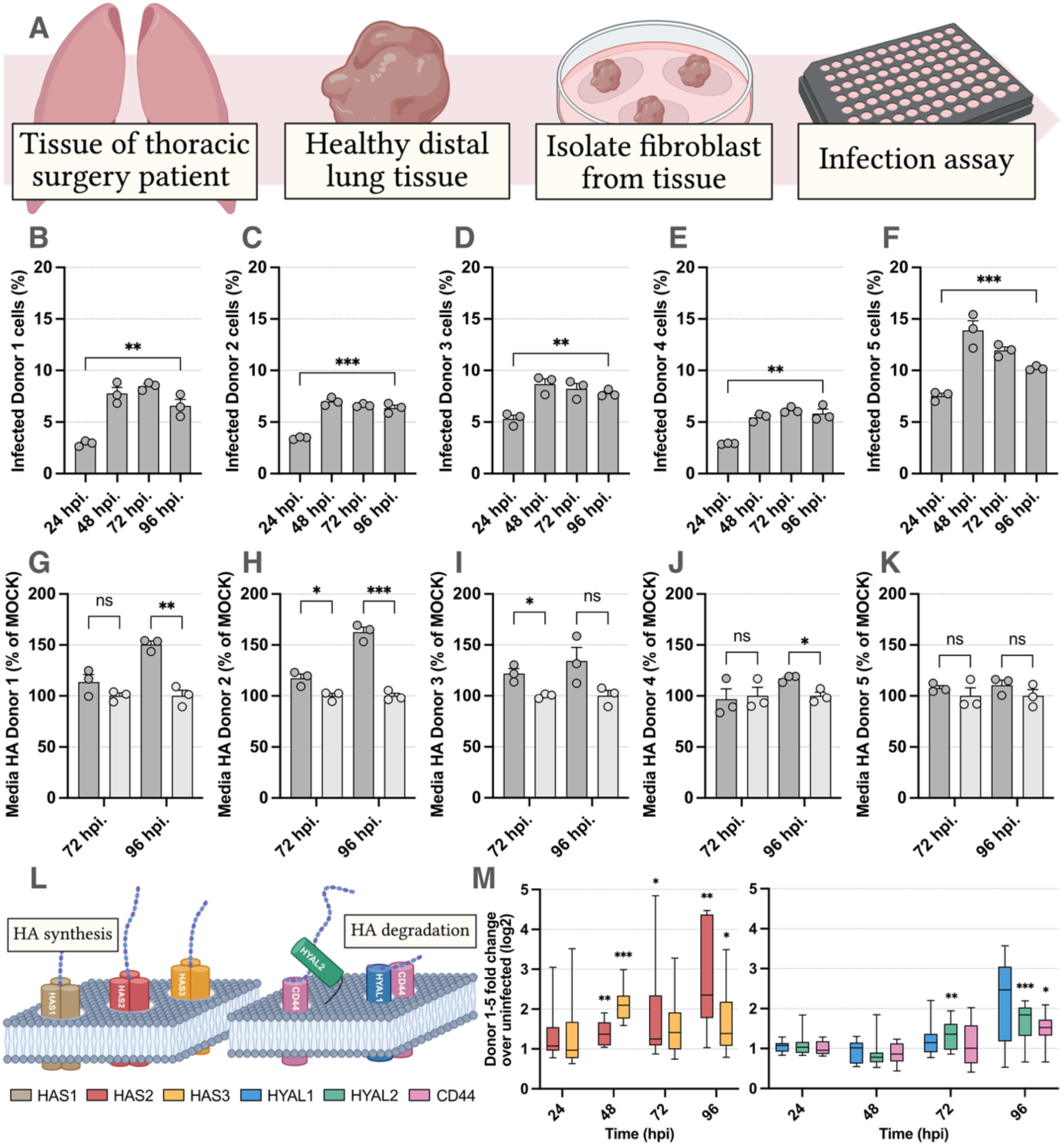
Primary human lung fibroblasts respond to Puumala orthohantavirus (PUUV) infection with increased hyaluronan (HA) production and dysregulation of genes involved in HA metabolism. (A) Schematic overview of primary human lung fibroblast isolation from healthy lung tissue obtained during thoracic surgery. (B–F) PUUV infection kinetics in primary lung fibroblasts from five individual donors, quantified at the indicated time points by immunostaining for PUUV nucleocapsid protein with nuclear counterstaining. Each panel represents fibroblasts derived from a single donor. Statistical significance calculated by unpaired t-test (*P < 0.05, **P < 0.01 and ***P < 0.001; ns, not significant). (G–K) HA concentrations in culture supernatants collected from PUUV-infected primary lung fibroblasts at 72 and 96 h post infection, measured by ELISA and expressed as percentage of uninfected controls. Statistical significance was assessed using multiple unpaired t-tests with Holm–Šídák correction for multiple comparisons (*P < 0.05, **P < 0.01, ***P < 0.001; ns, not significant). (L–M) Expression of genes involved in HA synthesis (*HAS1–3*), degradation (*HYAL1–2*), and signaling (*CD44*), determined by quantitative PCR, normalized to *β-actin*, and shown as fold change relative to uninfected controls. Data represents two independent samples per donor. Results are displayed as box-and-whisker plots, where the box indicates the 25th to 75th percentiles (interquartile range), the line within the box represents the median, and the whiskers denote the minimum and maximum values. (L) Schematic overview of genes involved in the HA synthesis, degradation and signaling. In panel (M), data from all donors are combined to illustrate overall trends. Statistical significance was assessed using paired t-tests, comparing the expression values to the respective time-matched uninfected controls (*P < 0.05, **P < 0.01, ***P < 0.001).

All donors supported productive infection and displayed comparable infection dynamics, characterized by an initial increase in infection levels followed by a gradual decline (Fig. 5B-F). Peak infection occurred between 48 and 72 hpi and ranged from approximately 8% (donor 4) to 15% (donor 5), indicating moderate variation in viral susceptibility between individuals. We next quantified HA levels in culture media from infected primary HLFs and time-matched uninfected controls. HA accumulation was observed predominantly at the later time points of infection, but with marked donors-to-donor variability. Four donors (donor 1-4) exhibited increased HA levels, reaching 110-160% relative to time-matched controls (Fig. 5G-J). In contrast, fibroblasts from donor 5 showed no detectable HA accumulation despite exhibiting the highest infection levels (Fig. 5K).

To further characterize the molecular basis of this heterogeneity, we quantified expression of HA-metabolic genes, including *HAS1-3*, *HYAL1-2* and *CD44* (Fig. 5L). *HAS1* expression was undetectable in the primary HLFs from all donors. Overall, the fibroblasts displayed a transcriptional response similar to that observed in lung-derived cell lines, characterized by early induction of *HAS2* and *HAS3* followed by delayed upregulation of *HYAL2* and CD44 (Fig. 5M). However, the magnitude and temporal dynamics of this response varied substantially between donors.

Together, these findings demonstrate substantial inter-individual heterogeneity in PUUV-induced HA dysregulation in primary lung fibroblasts, consistent with the variability observed among patients. Differences in HA accumulation were not explained by viral load alone, indicating that host-specific cellular factors influence the extent of pulmonary HA dysregulation during PUUV infection.

## DISCUSSION

In this study, we identify HA dysregulation as a previously unrecognized feature of PUUV infection. We demonstrate that systemic HA levels are transiently elevated during acute disease and correlate with clinical disease severity, and that HA accumulates extensively in the lungs of PUUV infected patients. Complementary *in vitro* experiments show that PUUV infection perturbs HA metabolism in human lung cells, identifying alveolar epithelial cells and lung fibroblasts as key contributors to infection-induced HA accumulation. Together, these findings implicate HA dysregulation as a component of PUUV-associated pathology and provide new insight into mechanisms underlying pulmonary involvement during hantavirus infections.

Our patient data demonstrates that systemic HA concentrations are markedly elevated during the acute phase of PUUV infection, increase in proportion to disease severity, and return to levels comparable to healthy controls during convalescence. This dynamic pattern suggests that circulating HA reflects active disease processes rather than baseline inter-individual variation. Similar transient increases in endothelial glycocalyx degradation products in plasma, including heparan sulphate, chondroitin sulphate, and HA, have been reported in hantavirus-induced HFRS and are associated with increased disease severity [26, 27]. Early inflammatory responses are a hallmark of hantavirus infections, and cytokine-driven glycocalyx degradation is a recognized feature of the disease pathogenesis [28, 29]. As HA is a major structural component of the glycocalyx, inflammatory injury to the vascular endothelium is expected to alter HA abundance and turnover. Importantly, accumulating evidence indicates that HA released during inflammatory injury is not merely a passive by-product but can actively exacerbate disease. In particular, low-molecular weight-HA promotes endothelial injury and barrier dysfunction in lung microvascular endothelial cells [30]. Together, these observations indicate that systemic HA is involved in inflammatory vascular injury and raise the possibility that HA may also contribute to pulmonary manifestations during PUUV infection.

Consistent with this notion, our histological analyses revealed extensive HA accumulation in the lungs of deceased PUUV infected patients, characterized by thickened alveolar walls and alveolar spaces partly or fully filled with HA-rich exudate. Progressive loss of alveolar epithelial-associated HA accompanied increasing tissue damage together with marked infiltration of CD45⁺ leukocytes, indicating substantial inflammatory cell recruitment. These findings define a pulmonary microenvironment characterized by extracellular matrix remodeling, epithelial disruption, and immune activation. Elevated HA in BALF further correlated with increased lung involvement. Notably, the HA detected in BALF was predominantly high-molecular weight, suggesting that accumulation primarily reflects increased synthesis or impaired clearance rather than extensive fragmentation at the time points examined.

Similar pathological features have been described in other severe respiratory viral infections. In COVID-19, extensive HA-rich exudates accumulate in the alveoli and are associated with alveolar wall thickening, impaired gas exchange, and progression to irreversible lung damage in severe cases [17, 19]. Similarly, respiratory syncytial virus and H1N1 influenza virus infections induce excessive pulmonary HA deposition and redistribution of subepithelial HA in murine models [20, 21]. Elevated HA levels in bronchoalveolar lavage fluid during influenza infection of mice parallels our observations in PUUV-infected patients [21]. Collectively, these findings place PUUV infection within a broader paradigm in which pulmonary HA accumulation is a recurring feature of severe viral lung disease.

A growing body of evidence indicates that degradation of the alveolar epithelial glycocalyx is a key pathological response to lung injury [31–34]. Glycocalyx disruption can promote increased HA synthesis and formation of large, cross-linked HA matrices associated with several pathological consequences. In the context of hantavirus, our observation of reduced epithelial-associated HA in lung tissue together with elevated HA levels in bronchoalveolar lavage fluid, particularly in patients with more severe lung involvement, support the notion that epithelial glycocalyx disruption and secondary HA overproduction contribute to pulmonary pathology. While the precise functional consequences of this HA accumulation remain to be determined, these findings suggest that HA is an active component of the local virus-induced inflammatory response in the lung.

Consistent with this potential role for HA in lung pathology, pulmonary involvement in PUUV infection has been documented in several clinical studies, with a substantial proportion of patients developing respiratory symptoms, radiographic infiltrates, and impaired oxygenation. Circulating glycocalyx degradation products in HFRS have been reported to correlate not only with overall disease severity but also with need for oxygen supplementation [27]. Widespread endothelial injury is therefore thought to increase vascular permeability, allowing glycocalyx components to enter the alveolar space and impair gas exchange [27]. Our data extends this model by demonstrating that PUUV infection also induces HA production in lung-resident epithelial cells and fibroblasts. Local HA synthesis may therefore act together with vascular leakage to amplify HA accumulation within the alveolar space. Given HA’s strong water-retaining capacity, such accumulation could promote fluid retention and oedema formation, providing a plausible mechanistic contribution to respiratory impairment in severe PUUV infection, including the minority of cases progressing to ARDS [9–11]. Although direct evidence in hantavirus HPS is currently lacking, similar mechanisms may operate during infections where pulmonary capillary leak and respiratory failure dominate pathology. Future studies examining HA dynamics in HPS will be important to determine whether HA represents a shared pathogenic mediator across the hantavirus spectrum.

Our *in vitro* data further demonstrate that PUUV-induced HA dysregulation is cell-type specific, with lung fibroblasts and alveolar epithelial cells representing the primary contributors to HA accumulation. Across cell types, PUUV infection induced early upregulation of HA synthases followed by delayed induction of hyaluronidases and CD44, creating a transient imbalance favoring HA accumulation. This temporal pattern correlated with the peak in extracellular HA levels observed during infection. Previous studies have shown that infection-induced inflammatory cytokines regulate both HA synthesis and degradation [35–37], supporting the idea that infection-induced inflammatory signaling influences HA metabolism. Notably, although a similar transcriptional response was observed across multiple cell types, the magnitude of HA accumulation varied, suggesting that additional cell-intrinsic factors modulate the outcome. HA dysregulation occurred independently of STAT1-mediated antiviral signaling, suggesting regulation primarily through infection-induced inflammatory pathways rather than classical antiviral restriction mechanisms; however, whether this response requires active viral replication or is primarily driven by secondary inflammatory signaling remains to be determined. The absence of detectable HA accumulation in lung microvascular endothelial cells *in vitro* further suggests that endothelial-associated HA alterations *in vivo* may predominantly reflect glycocalyx shedding rather than *de novo* synthesis, although this warrants further investigation.

Pronounced donor-to-donor variability in HA responses among primary human lung fibroblasts highlights an additional layer of host-dependent regulation. Despite comparable infection levels, fibroblasts from different donors exhibited divergent HA accumulation profiles, indicating that HA dysregulation is shaped by host-specific, cell-intrinsic factors rather than viral load alone. This heterogeneity mirrors the inter-individual variation in pulmonary involvement observed among PUUV infected patients and suggests that host determinants of HA metabolism may influence disease severity. While primary fibroblasts represent only one lung cell population, our data demonstrates that multiple lung-resident cell types contribute to HA production, implying that pulmonary HA accumulation *in vivo* likely reflects the combined response of several cellular compartments.

A major strength of this study is the integration of patient-derived material with mechanistic *in vitro* models, bridging clinical observations and cellular processes. The use of multiple lung cell types, together with primary human fibroblasts, enabled identification of both cell-type-specific and donor-dependent HA responses, while inclusion of consecutive time points allowed capture of dynamic changes in HA metabolism throughout infection. At the same time, our findings highlight the importance of future studies employing more complex experimental systems. *In vitro* monocultures do not fully recapitulate the infected lung environment, where endothelial cells, immune cells, and intercellular crosstalk likely shape HA dynamics. Future studies employing co-culture systems, advanced lung models, and targeted modulation of HA pathways will be essential for defining the functional contribution of HA to PUUV-induced lung pathology. In addition, pulmonary analyses were performed in a relatively small subcohort of patients, reflecting the invasive nature of bronchoscopy during acute infection. Larger studies will therefore be required to further validate the association between pulmonary HA accumulation and lung involvement.

Collectively, these findings support a model in which HA dysregulation represents a converging mechanism linking systemic vascular injury and local pulmonary pathology during PUUV infection. While hantavirus disease has traditionally been conceptualized primarily as a disorder of endothelial dysfunction and capillary leak, our data suggest that extracellular matrix remodeling within the lung parenchyma is an additional and potentially underappreciated component of disease pathogenesis. By demonstrating both vascular-associated HA elevation and local HA overproduction in infected lung cells, this study expands the current paradigm of hantavirus pathology beyond glycocalyx degradation alone. Although our data establishes strong clinical associations and mechanistic plausibility, direct evidence demonstrating that HA accumulation drives pulmonary dysfunction in vivo remains to be established.

From a clinical perspective, HA may represent both a biomarker and a potential therapeutic target in hantavirus infections. Elevated plasma HA could aid in identifying patients at risk of severe disease, while interventions aimed at modulating HA synthesis, degradation, or signaling may alleviate pulmonary oedema and inflammation. Given the similarities between PUUV infection and other HA-associated lung pathologies, such approaches may have broader relevance for viral-induced lung injury beyond hantaviruses.

In summary, we demonstrate that PUUV infection disrupts HA homeostasis in the lung, that HA accumulation correlates with disease severity, and that host-specific cellular responses shape this process. These findings advance our understanding of hantavirus-induced pulmonary pathology and identify HA metabolism as a novel axis in hantavirus pathogenesis.

## Acknowledgements

We sincerely thank all patients and control participants for taking part in this study. We also appreciate the valuable support provided by the staff of the Departments of Infectious Diseases and Respiratory Medicine & Allergy at University Hospital, Umeå. This study was supported by Kempestiftelserna (grant no. JCSMK23-0108 to A.L), Heart-Lung Foundation (20100192, 20150752, and 20170334 to C.A.), Emil and Wera Cornell’s foundation (A.L), Carl Bennet AB (A.L), County Council of Västerbotten, Umeå University (RV-457671 to C.A., RV-1035833 & RV-1028764 to T.T.), the Fundraising Foundation for Medical Research, Umeå University (978018 to A.L). Apart from funding, the sponsors were not involved in performing the present study.

Figure 1A, 3A, 4A, 5A and 5L were created in BioRender, Wennemo, A. (2026):

https://BioRender.com/4ey80bg, https://BioRender.com/3x19jm4,
https://BioRender.com/h372369, https://BioRender.com/yvhdgcc.

