## Supplemental information for "Puumala orthohantavirus dysregulates hyaluronan metabolism in lung cells and correlates with disease severity and lung impairment"

### SUPPLEMENTARY INFORMATION

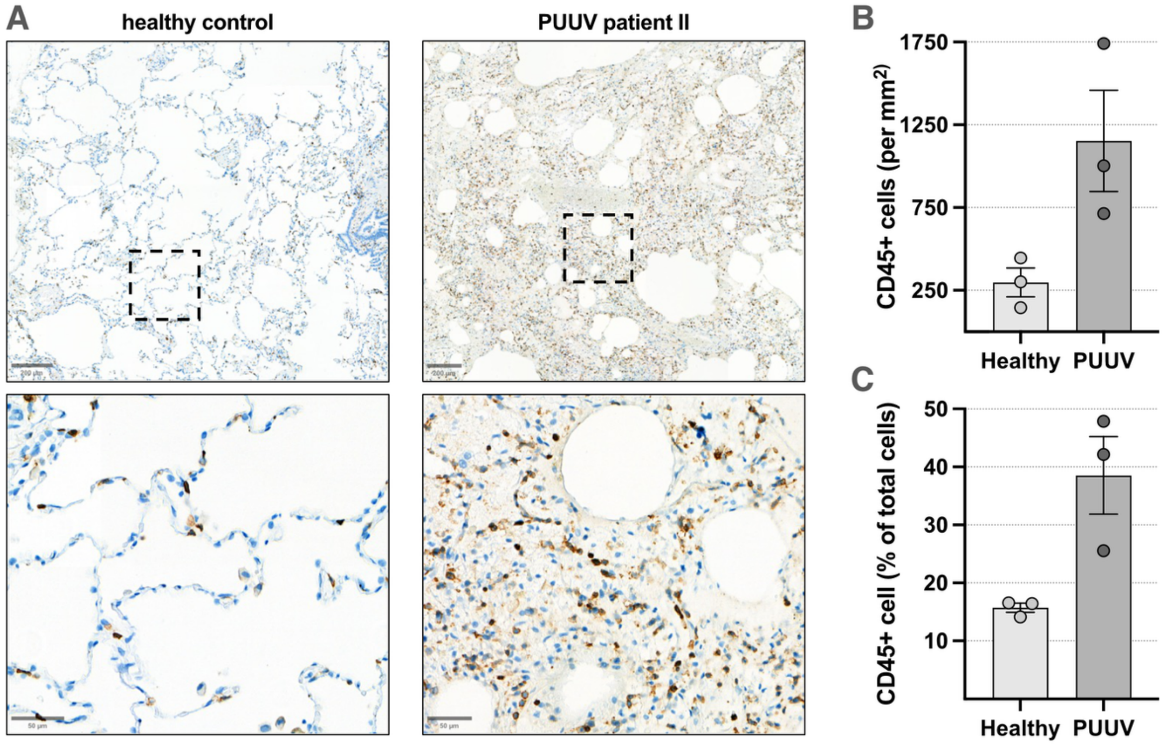


**Supplementary figure 1. Leukocyte infiltration increases in lung tissue of Puumala orthohantavirus (PUUV) infected patients.** (A) Lung tissue section from healthy controls (n = 3) and deceased PUUV infected patients (n = 3) were stained for CD45 and counterstained with hematoxylin and bluing reagent to visualize nuclei. The upper panel shows representative overview images of healthy and PUUV-infected lung tissue. Scale bar, 200 µm. The lower panel displays higher-magnification images of indicated regions from the same sections, highlighting differences in cell density and tissue morphology. Scale bar, 50µm. (B-C) Quantification of $CD45^+$ cells in healthy and PUUV infected lung tissue (B) represented as number of $CD45^+$ cells per $mm^2$ of tissue and (C) proportion of $CD45^+$ cells of total cell count. For quantification, ten random 1 $mm^2$ squares were positioned on the tissue for quantification of $CD45^+$ and total cell count. Each circle represents the average of the ten random subsections for each tissue section.

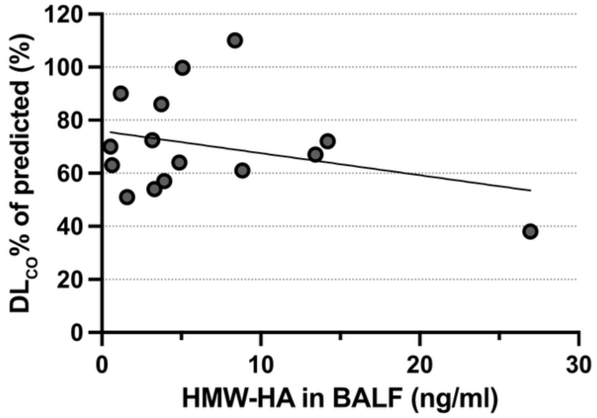


**Supplementary figure 2. Association between hyaluronan (HA) and predicted lung diffusion capacity in Puumala orthohantavirus (PUUV) infected patients.** Simple linear regression analysis showing predicted diffusion capacity ($DL_{CO}$% predicted) in relation to the high-molecular weight (HMW) HA in the bronchioalveolar lavage fluid (BALF) during the acute phase PUUV infection. Linear regression revealed a non-significant negative association (β = -0.830, $R^2$ = 0.093, $P$ = 0.268).

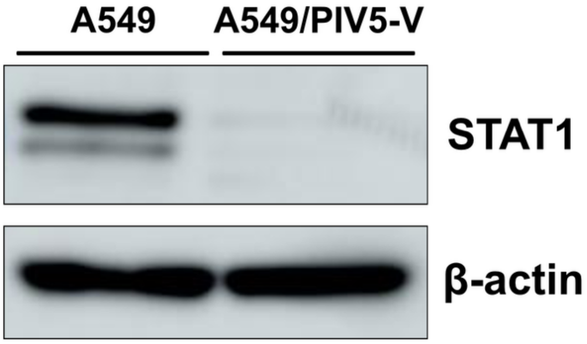


**Supplementary figure 3. STAT1 expression in A549 and A549/PIV5-V cells.** Western blot analysis of STAT1 expression in A549 and A549/PIV5-V cell lysate. Total protein of cell lysate was separated on an SDS-PAGE and transferred to a PVDF membrane. The membrane was probed with anti-STAT1 (1:500) and β-actin (1:1000) primary antibodies followed by an HRP-conjugated secondary antibody. Bands corresponding to STAT1 (88 kDa) and β-actin (44 kDa) were detected using chemiluminescence. β-actin served as a loading control.
